# Post-pandemic modeling of COVID-19: Waning immunity determines recurrence frequency

**DOI:** 10.1101/2023.01.16.23284640

**Authors:** D Calvetti, E Somersalo

## Abstract

There are many factors in the current phase of the COVID-19 pandemic that signal the need for new modeling ideas. In fact, most traditional infectious disease models do not address adequately the waning immunity, in particular as new emerging variants have been able to brake the immune shield acquired either by previous infection by a different strain of the virus, or by inoculation of vaccines not effective for the current variant. Furthermore, in a post-pandemic landscape in which reporting is no longer a default, it is impossible to have reliable quantitative data at the population level. Our contribution to COVID-19 post-pandemic modeling is a simple mathematical predictive model along the age-distributed population framework, that can take into account the waning immunity in a transparent and easily controllable manner. Numerical simulations show that under static conditions, the model produces periodic solutions that are qualitatively similar to the reported data, with the period determined by the immunity waning profile. Evidence from the mathematical model indicates that the immunity dynamics is the main factor in the recurrence of infection spikes, however, irregular perturbation of the transmission rate, due to either mutations of the pathogen or human behavior, may result in suppression of recurrent spikes, and irregular time intervals between consecutive peaks. The spike amplitudes are sensitive to the transmission rate and vaccination strategies, but also to the skewness of the profile describing the waning immunity, suggesting that these factors should be taken into consideration when making predictions about future outbreaks.

## 1 Introduction

The outbreak of the COVID-19 pandemic in early 2020 dramatically altered the landscape of public health, clinical and epidemiological research, and changed profoundly the public perception of the risks of the disease. While the level of immunity of the population to the SARS-CoV-2 virus has been boosted by the production of effective vaccines and widespread vaccination campaigns, expectations about the long term fate of the pandemic have changed following the periodic discovery of new pathogen mutations with varying virulence and transmissibility that can elude both vaccine-based and disease-acquired immunity. Other important factors that need to be taken into account when making predictions are that the evolution of the public perception about the severity of the infection and a growing level of mitigation fatigue have lowered the level of alertness and protection against the disease. In several countries, in part because of social and economic pressure, COVID-19 has been downgraded from a health crisis to a health concern, and the almost ubiquitous removal of the obligation to report test results, paired with widely available self tests have made almost all infection data at the population level obsolete and unreliable. These changes in the pandemic landscape pose new challenges to predictive mathematical models. In the current post-pandemic period, there are several questions in need of fast answers, including how the old models should be modified to be useful in the current phase. Another question is what are the relevant questions that mathematical models can answer, and what kind of data, if any, should be used to estimate model parameters and validate model predictions. A more fundamental question is if mathematical models still have a predictive role in COVID-19 responses, whether the focus should shift towards more qualitative goals. In other words, what should the post-pandemic modeling paradigm be? This paper begins addressing these questions by defining new goals for mathematical models and presenting some examples of predictive models adhering to the changing paradigm.

As the obligation to report new infections makes the reliability of the available data quite questionable, it becomes necessary to look for data sources that transcend voluntary reporting. Possible sources of information that do not require an active role of the population include the number of hospitalizations due to the disease, COVID-related death rates, and measurements of viral load in sewage. All of these data types lack direct information about the prevalence of new infections in the population, thus estimates of detailed model parameters such as transmission rate based on them could be highly unreliable and uncertain. Although numerically unreliable, the information that could be extracted from these data is whether the infections are increasing or decreasing, and when the disease is expected to peak. Predictions about the timing of the next infection peak, and its width could be used not only to assess the reliability of a model, but also for streamlining public health operations, e.g., planning the staffing of hospitals and estimating the number of beds to be allocated for COVID-19 patients.

From the very onset of COVID-19, efforts to understand various aspects of the transmission at individual and population level have generated a huge corpus of modeling literature, see, e.g. [7] for a meta-analysis of the approaches and objectives of the research. Most contributions in the literature do not address the novel feature characterizing the current phase of the pandemic, namely the role of interval of time before immunity wanes and the arrival of new variants capable of eluding the immunity provided by vaccination or by exposure to a previous variant of the pathogen. However, traditional epidemiological models based on ordinary differential equations such as the classical SIR model can be modified to take into account the loss of immunity, in particular, to model endemic diseases. The most straightforward modification is to include a flux from the recovered compartment back to the susceptible compartment (SIRS model), see, e.g., [1]. More sophisticated models augment the SIRS model or its variants [9, 11] by adding a waning immunity compartment (W) accounting for those individuals who are either losing the immunity, or alternatively, may get an immunity boost through a new exposure to the virus. For a review of classical SIRWS models for endemic diseases such as pertussis, we refer to, e.g., [3, 5, 15, 16]. These SIRWS models have been recently revisited in the context of COVID-19, see [14, 19, 22]. A remarkable property of these models is their ability to generate periodic solutions and recurrent infection spikes even in short time intervals, reproducing the recurring waves of infections shown in COVID-19 news outlets, through a mechanism describing the arrival of new variants and the gradual loss of acquired immunity over time.

In the current phase of COVID-19, the recurrence of infection waves continues to draw attention from mathematical modelers and public health officials alike. In light of the current reporting situation, overly complicated models depending on large number of parameters with detailed hypothetical transmission mechanisms may not be practical due to the lack of reliable data allowing the model validation. The present article proposes a conceptually simple compartment model addressing waning immunity in terms of an easily interpretable profile describing the time evolution from immune to vulnerable. The model follows an age-distributed population model formalism based on Leslie matrices describing the evolution of the immunity level. Computed experiments demonstrate that the immunity profile characteristic to the disease’s current phase is the key factor behind the infection recurrence. Other factors, including human behavior or infectivity of the pathogen, while important for determining the amplitude of the spikes, seem to have a secondary role, albeit not being without significance. The rest of the paper is organized as follows. In the next section we introduce the mathematical model and explain the formalism utilized. Section 3 is dedicated to understanding the model parameters in terms of quantities that can be related to the available data. The significance of the values of the model parameters in determining the type of behavior of the pandemic, in particular in relation to whether or not recurrent waves can be expected, is discussed at length. Model simulations with different parameter values are used to support the proposed causal relation. A discussion of the findings and some conclusions about the refocused role of mathematical models in the post-pandemic era presented in section 4.

## 2 Model

In this section, we develop a simple, discrete-time deterministic infection model with an advection structure, resembling classical age-structured population models based on the Leslie matrix formalism [12, 10]. We begin by describing a version of the model that does not include vaccination, then later explain how that can be added.

Let *N* = *N* (*t*) denote the total population size at time *t*, where time is discretized in units such as one day and assumed to take on integer values, with the understanding that the time units can be modified as needed.

We begin by subdividing the population into two cohorts, the *susceptible* (S) and the *infected* (I). The sizes of these cohorts at time *t* are denoted by *S*(*t*) and *I*(*t*), respectively, and

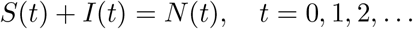

Consider first the susceptible population S. The underlying hypothesis in the basic model is that immunity, acquired through disease exposure, wanes as time from the last recovery increases. Therefore, we divide the susceptible cohort into *L* subgroups, S_1_, …, S_*L*_. If *s*_*ℓ*_(*t*) denotes the number of individuals in the _*ℓ*_th subgroup S_*ℓ*_ at time *t*, or briefly, the size of S_*ℓ*_, where

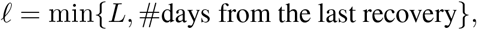

then

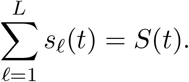

Each subgroup is characterized by its *vulnerability* to infection, intended as degree of lack of immunity. We denote the vulnerability of S_*ℓ*_ by *w*_*ℓ*_, with

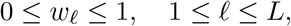

where *w*_*ℓ*_ = 0 means full immunity to the disease, and *w*_*ℓ*_ = 1 means a total lack of immunity. For simplicity, the vulnerabilities are not time dependent. We assume that the vulnerability is a non-decreasing function of _*ℓ*_, that is, *w*_*ℓ*_ *≤ w*_*ℓ*+1_. With this interpretation, *L* can be seen as an estimated upper bound for the duration of the immunity protection.

In our model, we postulate the following guiding principles:

1. The number of individuals in S_*ℓ*_ becoming infected in a unit time interval [*t, t* + 1] is proportional to
  a. the vulnerability *w*_*ℓ*_ of S_*ℓ*_,
  b. the size *s*_*ℓ*_ (*t*) of S_*ℓ*_,
  c. the total fraction of infective individuals in the entire population.
2. The increase of vulnerability is described through advection with a time step. When the last vulnerability class S_*L*_ is reached, the vulnerability no longer increases with time.

According to these assumptions, if at time *t* individuals from the subgroup S_*ℓ*_ get infected, they will be removed from the subgroup. If

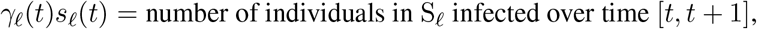

then

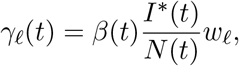

where *β*(*t*) is the transmission rate, or pairwise infectious contact rate, at time *t*, and *I*^*∗*^(*t*) is the size of the subpopulation of the cohort I that is infective, i.e., individuals with an active viral load. Here, we assume that *γ*_*ℓ*_ (*t*) *<* 1. Every individual in the group S_*ℓ, ℓ*_ *< L*, will be removed, either by joining the infected compartment, or by being advected to the next vulnerability class. Therefore, we conclude that

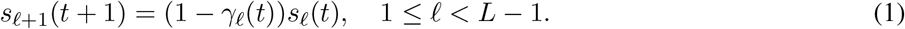

Since the last vulnerability class S_*L*_ contain those individuals who were already in the class and did not get infected, and those arriving from class S_*L−*1_, the formula for the update of its size becomes

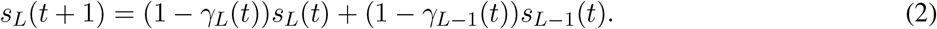

Before deriving the update formula for the size of the first class S_1_, we consider the infected cohort I and subdivide it into *K* subgroups, I_1_, …, I_*K*_. If we let *i*_*k*_(*t*) denote the number of individuals in the *k*th subgroup I_*k*_, then

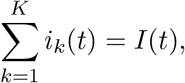

where

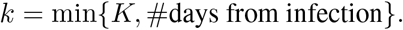

We set the value of *K* to the smallest number of time units after which infected individuals are no longer infective, so that the number of infective individuals at time *t* is given by

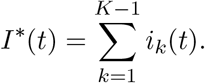

Individuals in each group I_*k*_ either recover, die, or stay infected. We introduce the *recovery rate η*_*k*_ and the *death rate µ*_*k*_ of the subgroup I_*k*_,

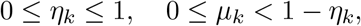

assuming that the rates are not time dependent. We postulate the following modeling principles.

1. The number of individuals in I_*k*_ that recover durung [*t, t* + 1] is proportional to
  a. the recovery rate *η*_*k*_ of I_*k*_,
  b. the size *i*_*k*_(*t*) of I_*k*_.
2. The number of individuals removed by death from each group I_*k*_ in the interval [*t, t* + 1] is proportional to
  a. the death rate *µ*_*k*_ of I_*k*_,
  b. the size *i*_*k*_(*t*) of I_*k*_.
3. All non-recovered surviving individuals in I_*k*_ *k < K*, are moved automatically to the next infected group I_*k*+1_.
4. Individuals in group I_*K*_ that do not recover or die remain in I_*K*_.

According to our assumptions, if the recovery and death rates are invariant in time, we have

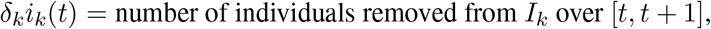

where

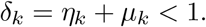

Therefore, the updating formula for the size of the infected subgroups is

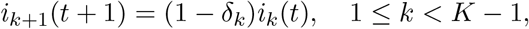

while for the last subgroup I_*K*_,

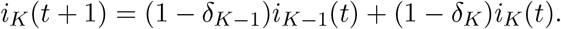

Following the assumption that recovered individuals have the highest level of immunity and move to the group S_1_, we have that

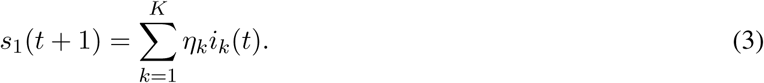

On the other hand, all susceptible individuals who acquire the disease over the time interval [*t, t* + 1] enter the infection group I_1_ hence

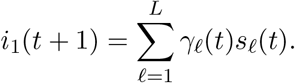

Figure 1 show a flow chart schematics of the model.

**Figure 1:**
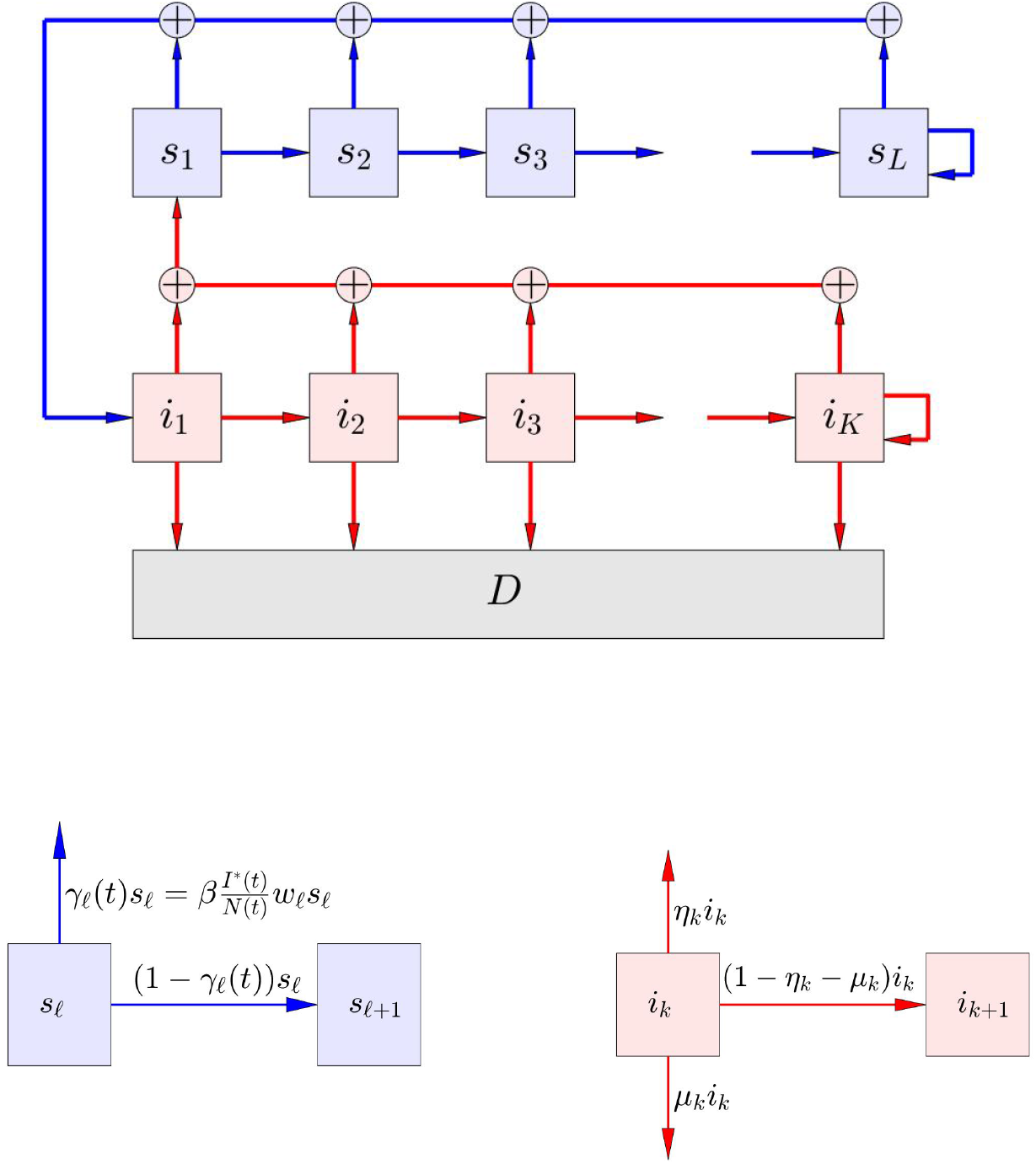
Schematic representation of the model. The blue boxes represent subsets of the susceptible population with a given level of immunity. As time goes by, the immunity level wanes, causing an advection flow towards the right for the part of the subpopulation that is not getting infected. The pink boxes represent subsets of the infected population, and the advection to the right indicates simply the time of the infection. The gray box indicates the deceased population due to the infection. The two inserts showing details of the schematics indicate the fluxes.

We are now ready to collect all updating formulas. We have

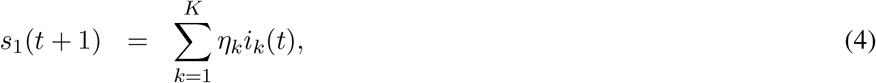

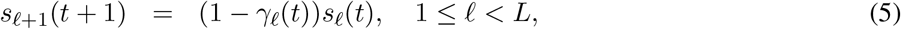

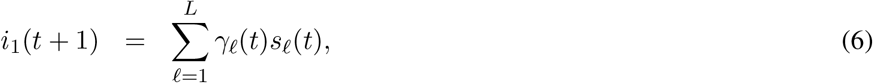

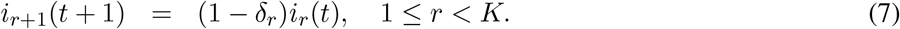

We introduce the matrices M_*ij*_, 1 ≤ *i, j* ≤ 2,

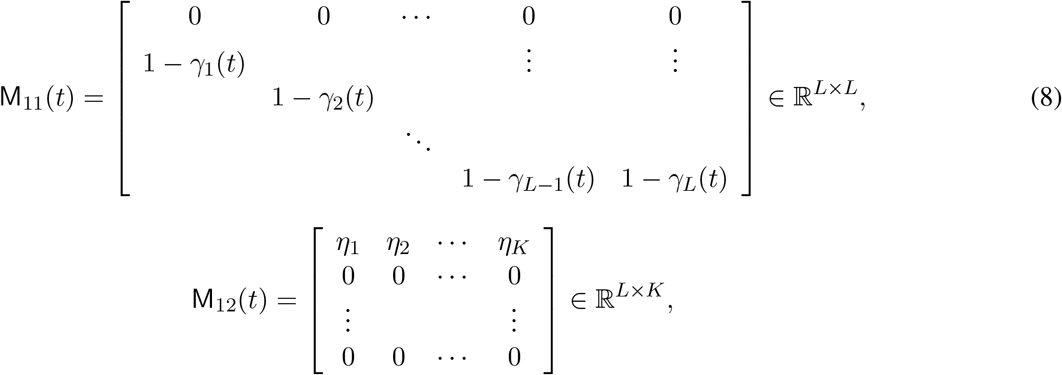

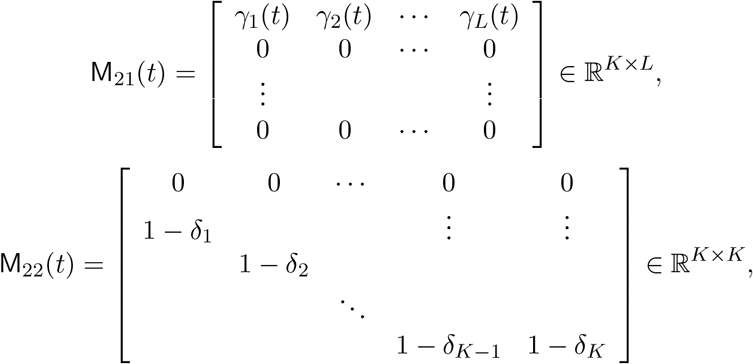

and define the Leslie matrix

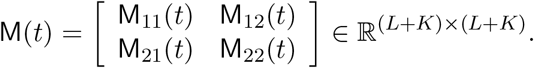

The governing equations of our discrete time dynamic model can then be expressed concisely as

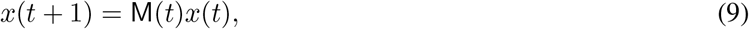

where

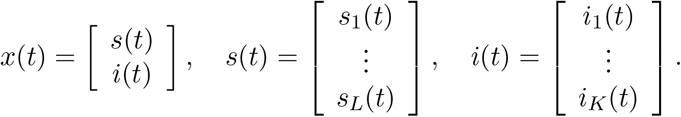

Observe that (9) is not a linear evolution model, because the factors *γ*_*R*_(*t*) depend on the state vector *x*(*t*), which can be emphasized by writing

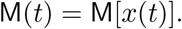

### 2.1 Vaccination

Until now, it was assumed that immunity can be acquired only through recovery from infection. The model can be modified to include vaccination by adding shunt pathways from S_*ℓ*_ to S_1_. More precisely, consider the compartment S_*ℓ*_ at time *t* and let

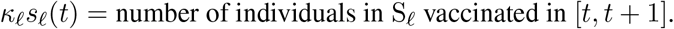

We account for the removal of vaccinated individuals S_*ℓ*_, by replacing (1) with

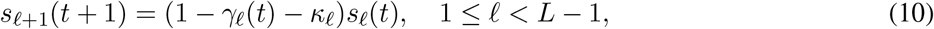

and (11) by

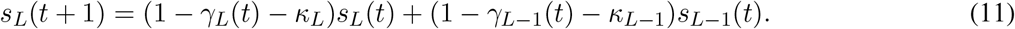

Moreover, since all vaccinated individuals move to S_1_, we change (3) to

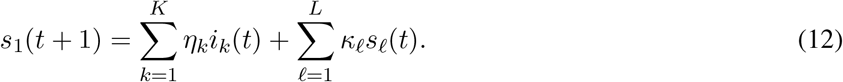

The addition of vaccination to the model requires only the modification of the matrix M_11_, replacing (8) by

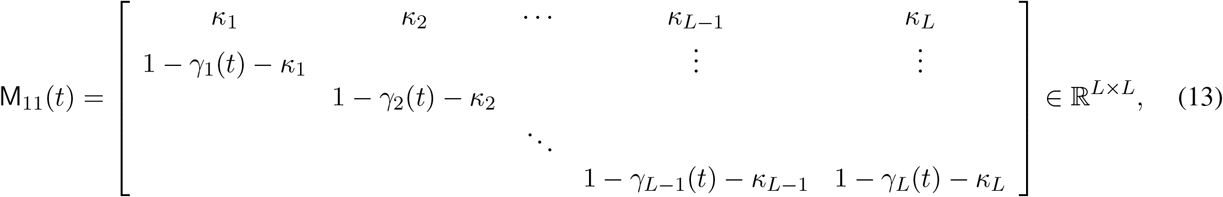

Observe that it is tacitly assumed that vaccination provides the same kind of immunity as the recovery from the disease, a hypothesis that may be challenged in the light of recent analysis [13]. Moreover, with the arrival of new variants of the virus, recovery from infection of one variant possibly may provide lower immunity against new ones. Mathematical models including different variants together with vaccination have been proposed in the literature, see, e.g., [2, 4]. In the literature there are models assuming different immunity levels depending on the age of the individuals [8]. The model that we are proposing here can be generalized to account for different strains of the virus and different types of immunity profiles through the additions of subpopulation models, at the cost of increased complexity.

## 3 Effects of the model parameters

One of the aims of this contribution is to use the proposed model to begin understanding the mechanisms that control the recurring infection waves observed since the onset of the pandemic. In this section, the oscillatory structure is discussed in the light of model simulations.

To set up the computed experiment, we need to assign values to the model parameters. We consider first the model without vaccination and assume a total population size of *N* = 10^6^. Since the focus here is on the infection dynamics, we set for simplicity *µ* = 0, so the population size remains fixed. The number of infected classes is set to *K* = 11, implying that nobody remains infective for more than 10 days, observing that the last class, I_*K*_ is not considered infective. To set the recovery rates, we assume that

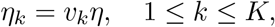

where 0 *< v*_1_ *< v*_2_ *<* … *< v*_*K*_ = 1, and *η* is the recovery rate after *K* days. We set *η* = 0.7 days^*−*1^, which amounts to assuming that after *K* days, recovery is expected in 1*/η ≈* 1.4 days. The increasing sequence of weights *v*_*k*_ follows a sigmoid, shown in the right panel of Figure 2.

**Figure 2:**
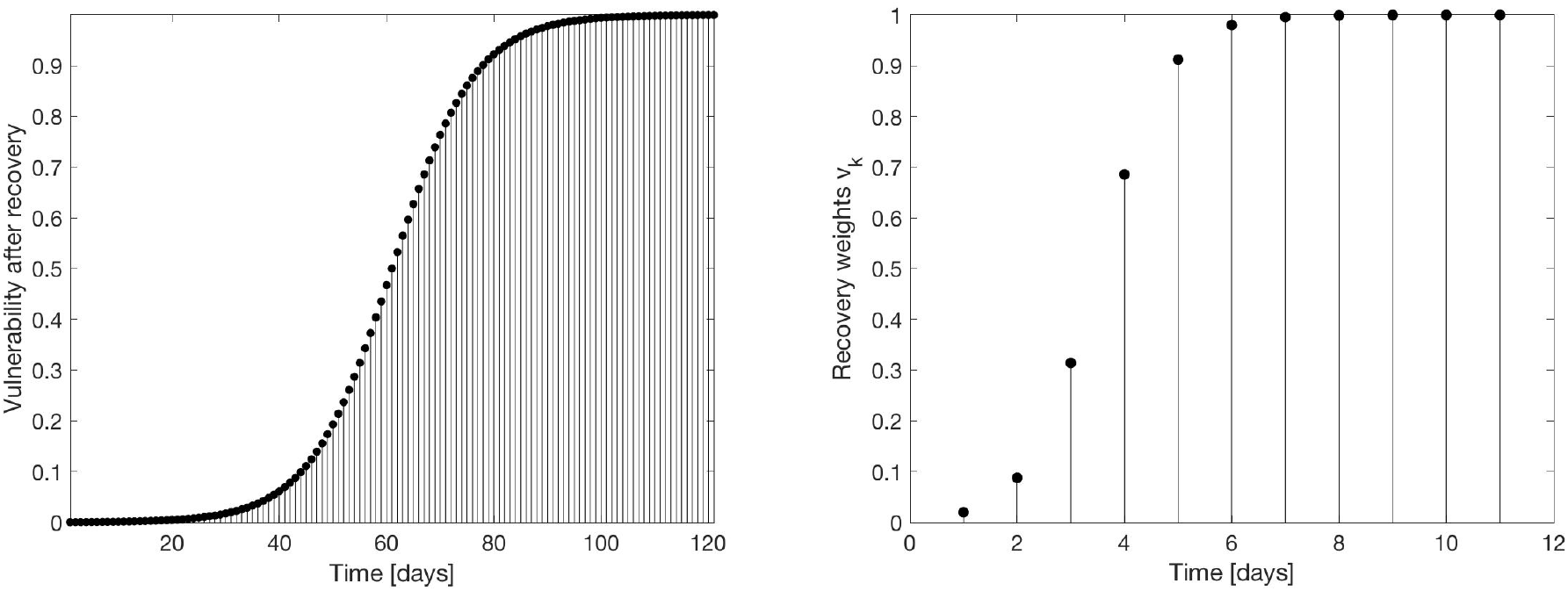
The vulnerability profile *w*_*k*_ with *L* = 120 days (left) and the recovery weights *v*_*k*_ (right) with *K* = 11. The vulnerability profile assumes that the immunity is completely lost after *L* days. The recovery profile indicates how the expected recovery increases as time passes.

### 3.1 Effect of the transmission rate

The first set of simulations analyze the role of the transmission rate *β*(*t*). Consider the susceptible population and assume that the immunity wanes over a period of *L* = 120 days. In the simulations, we use the vulnerability profile with 0 *< w*_1_ *<* … *< w*_*L*_ = 1 shown in Figure 2. The vulnerability profile is kept fixed, while the transmission rate *β* is varied. The role of seasonality in COVID-19 recurrence, previously in the literature is [6, 17] is considered also in the light of the current model.

#### Case 1

In the first computed example, we assume constant transmission rate, *β* = 0.4 days^*−*1^. In figure 3, the plot of new daily infections over a period of four years, shows a periodic behavior, with spikes repeating at regular intervals of approximately *L* = 120 days. We therefore conclude that with constant transmission rate, the periodicity is determined by the time it takes for immunity to be lost.

**Figure 3:**
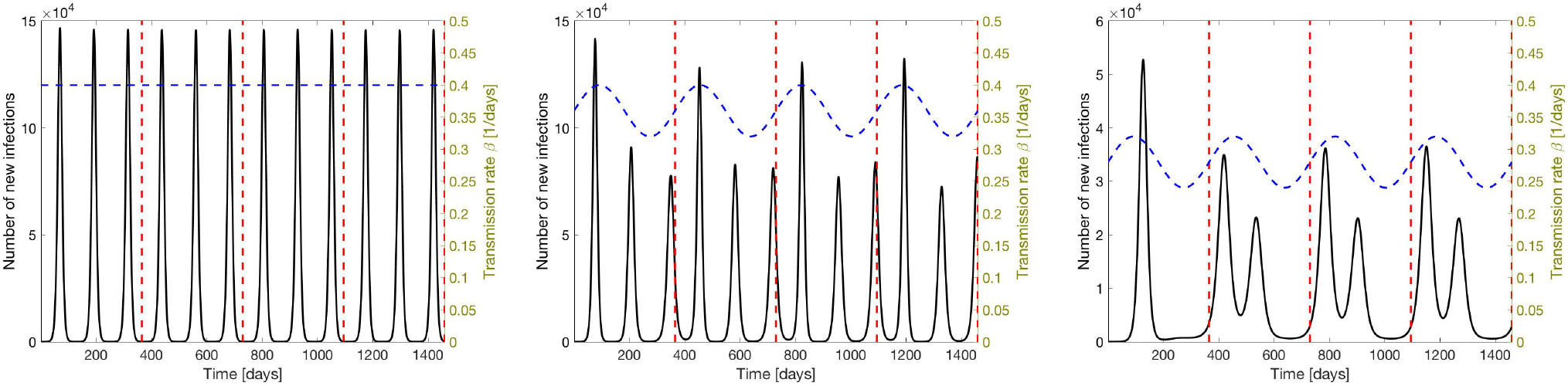
Simulated results showing the number of new daily cases (black curve). The simulation covers four years, each year marked by a vertical dashed red line. Immunity is assumed to wane in 120 days, or roughly one third of a year. In the left panel, the transmission rate (blue dashed curve) is held constant and set to *β* = 0.4 days^*−*1^, while in the middle and right panels, *β* is assumed to be time dependent with a sinusoidal behavior. Due to the lower mean transmission rate in the right panel the third annual peak is missing, because *β* is too low to create a rebound of the infections.

#### Case 2

This protocol simulates seasonal variations of the transmission rate *β*. To this end, we define the time dependent parameter with a periodicity of one year.

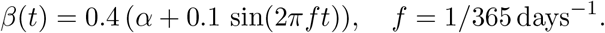

We set first *α* = 0.9, so that the maximum of *β* is the value used in Case 1. With this choice, the number of new infections retains the structure of three infection peaks per year, however, the amplitudes of the peaks vary corresponding to the variation of the infectivity. Next we set *α* = 0.7. In this case, *β*(*t*) during the third yearly peak is significantly lower than during the first two peaks, and while the yearly periodicity is retained, the third infection peak is missing. We therefore conclude that while the period of waning immunity still determines the periodic structure, seasonal changes may indeed lead to missing infection peaks.

#### Case 3

In the third protocol, we let the transmission rate vary randomly. More precisely, we generate a Gaussian random process *W*_*t*_ with Matèrn covariance with correlation length *λ* = 30 days (see, e.g., [23]), thus modeling medium-range changes in the pathogen infectivity as well as changing patterns of human behavior, e.g., adherence to social distancing and use of facial masks. The model for *β* is given by

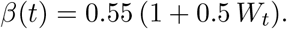

Figure 4 show three realizations of the new infection cases, together with the random time courses of the corresponding realizations of *β*. We observe that when the transmission rate becomes low, the spikes may not re-emerge. Furthermore, there is more variability in the spike separation. A long run with randomly varying transmission rate reveals that the spike separation tends to loose coherence.

**Figure 4:**
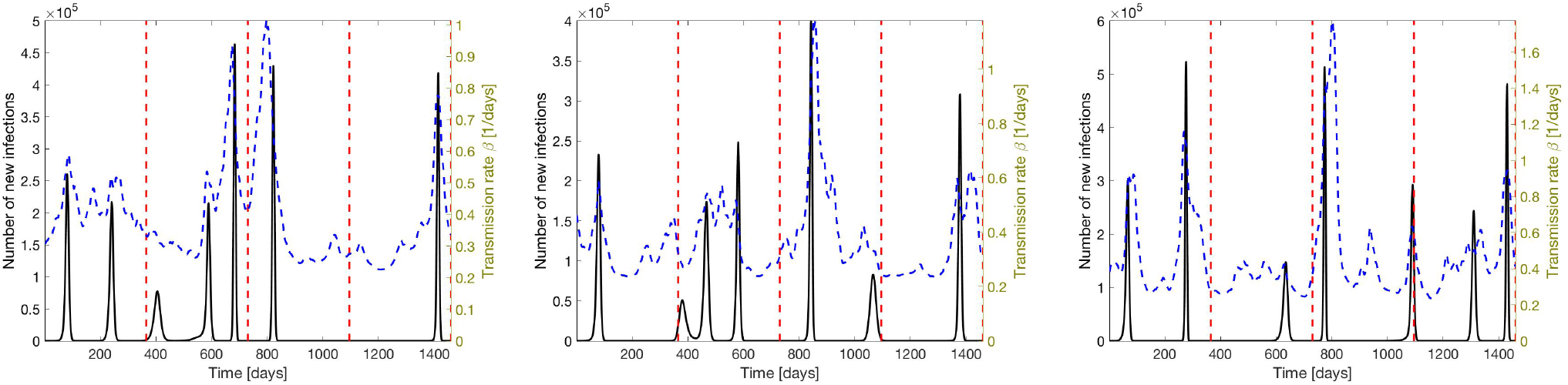
Three simulation corresponding to different realizations of the transmission rate *β*, modeled as a Gaussian process of Matèrn type, with correlation length *λ* = 30 days, simulating variations of contagiousness of the pathogen and the human behavior. The peaks appear near, but not exactly where they occurred with constant transmission rate *β*. Low transmission rate results in missing peaks. As time passes, the peak locations become less regular.

### 3.2 Vulnerability profile

The time constant defining the length of the waning immunity period was shown to be a decisive factor for the return rate of the infection waves. A legitimate question is how much the period depends on the actual vulnerability profile that was assumed to follow a sigmoidal behavior. To shed some light on this dependency, we run simulations with randomly varying vulnerability profile. More precisely, in our simulations, we represent the vulnerability profile as a convex combination of sigmoidal template profiles,

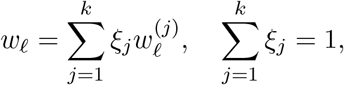

where the templates 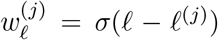 are sigmoidal functions with identical shape but shifted with respect to each other, with a ramp-up time equal to a fraction of the waning time *L*. In our numerical simulations, we use 11 template functions, each one having an approximate ramping time of 10 days. Figure 5 shows two realizations of the function *w*_*ℓ*_, both being convex combinations of three template functions with coefficients *ξ*_*j*_ as indicated in the plot.

**Figure 5:**
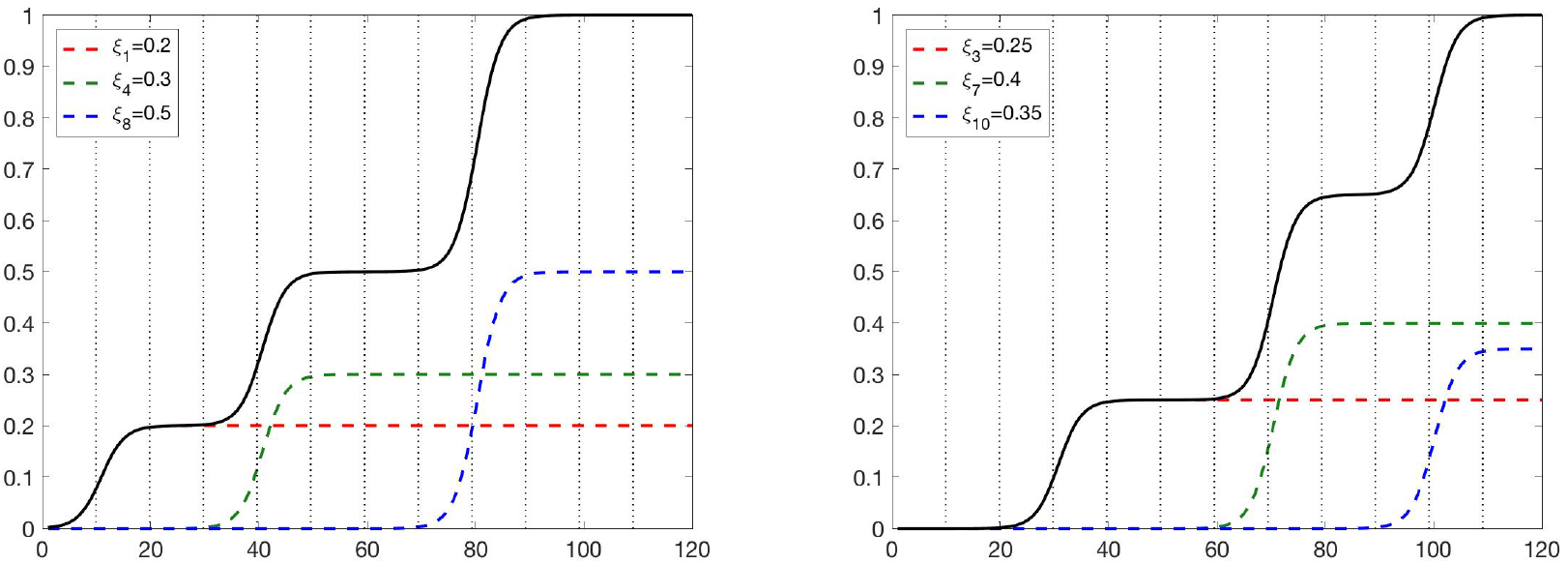
Two realizations of the function *w*_*ℓ*_ composed by three template functions, the non-zero weights *ξ*_*j*_ indicated in the figure. The vertical dotted lines indicate the positions of the 11 template functions used in the simulations.

Figure 6 shows four realizations of the vulnerability profiles with randomly drawn weights *ξ*_*j*_ and the corresponding computed new infection counts. Two characteristics of the outputs are worth highlighting. First, the repetition frequency of the infection spikes is fairly insensitive to the profile shape. The second observation is that in two of the simulations, the oscillations flatten out towards an asymptotic constant value. The figure shows also the cumulative new infection count over time. A natural question is, what feature of the vulnerability profile determines the asymptotic behavior of the solution.

**Figure 6:**
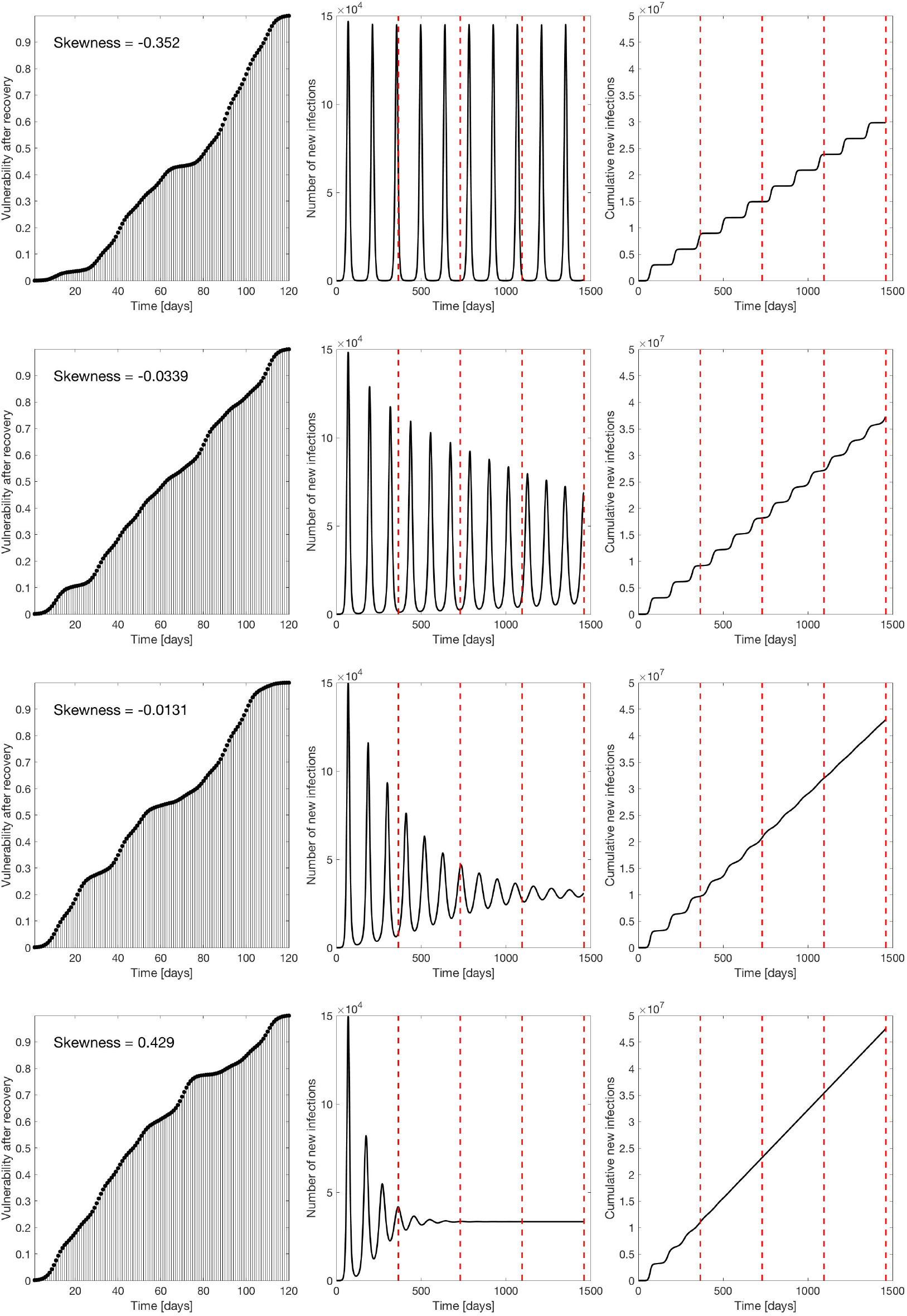
Four realizations of random vulnerability profiles (left column) and the corresponding simulated new infections (center column), and the cumulative new infections (right column). The skewness indicated in the plots refer to formula (14).

To answer this question, we normalize the vulnerability profile as a cumulative distribution of a probability density *π* over a unit interval [0, 1],

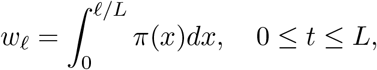

and compute the skewness of this density,

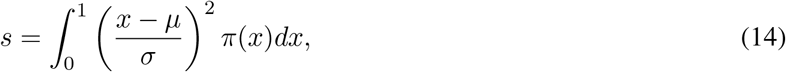

where *µ* and *σ* are the mean and the standard deviation of the density *π*. Simulations show that the damping of the oscillations depends strongly on the skewness: for *s <* 0, the oscillations show little or no damping, while for *s >* 0, the damping is significant. To demonstrate this effect, we generate a sample of *N* = 5 000 vulnerability profiles by drawing the coefficients *ξ*_*j*_ from uniform distribution over [0, 1] and normalizing them so that Σ*ξ*_*j*_ = 1. For each simulated profile, we generate the profile of new infections over a period of 4 years, and to indicate the damping, we compute the ratio of the amplitudes of last and the first infection spike. The plot in Figure 7 show a strong dependency of the damping factor of the skewness: Negative skewness means that the vulnerability grows slowly at the beginning of the time interval and strongly towards the end, and positive skewness implies the opposite, a strong growth at the beginning and slower growth towards the end, as suggested by the profiles in Figure 6.

**Figure 7:**
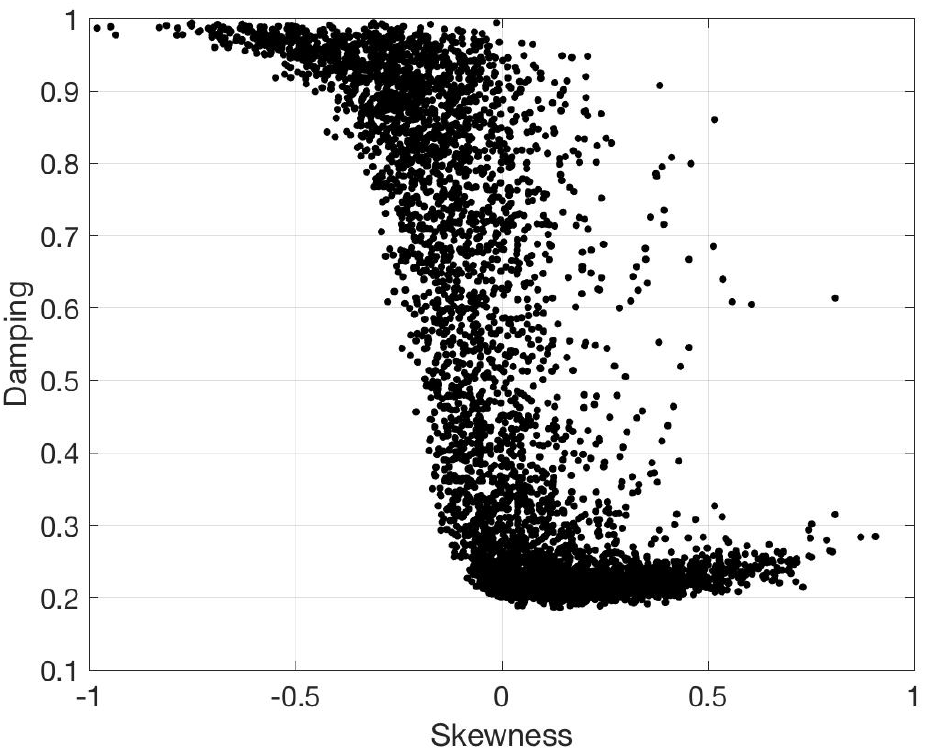
Skewness of the generating distribution of the vulnerability profile versus the damping factor, measured as the ratio between the last and the first spike amplitudes over a simulation period of four years.

### 3.3 Effect of vaccination

To understand the role of vaccination, we ran simulations based on the following two assumptions:

1. Recovered population is vaccinated not earlier than 90 days from the recovery from infection or previous vaccination;
2. The vaccination rate of the eligible individuals is constant among the subpopulations.

This is implemented by setting

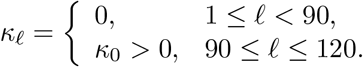

The simulations considered the three different models for the transmission rate discussed in Subsection 3.1. While not surprisingly, the vaccination lowers the number of newly infected, it turned out that unless the vaccination rate was not set at unrealistically high values (of the order *κ*_0_ ≈ 0.02, requiring a constant vaccination program of the order of 0.5% of the uninfected population per day), the qualitative behavior of the new infection count was retained. In the case of randomly fluctuating transmission rate, vaccination even with moderate rate values (*κ*_0_ ≈ 0.001) occasionally resulted in profiles of new infections in which some of the low level infection peaks essentially vanished. In summary, mathematical evidence from the model suggest that the effect of vaccination in the current setting is equivalent to an effective lower transmission rate.

## 4 Discussion and conclusions

In this paper we propose a simple model based on the paradigm of age-structured population models and use it to provide an intuitive and transparent explanation of the rebound mechanism of COVID-19 infection spikes. The model is controlled by two factors, the transmission rate and the waning immunity profile.

One of the questions that we have addressed is whether seasonal changes in the transmission rate are the primary explanation of the cyclic nature of the infection spikes. A recent study [6] found SARS-CoV-2 to be temperature and humidity sensitive, suggesting a seasonal nature of the disease. The study, however, was performed based on early COVID-19 data before the prevalence of new variants and significant reinfections, and the authors admit that the conclusions, based on the short observation period, may not be reliable. Another study [17] found that cold season contributes to the increase of COVID-19 cases. These findings are not in conflict with those in the present paper: increase in *β*, e.g., through indoors gatherings, amplifies the peaks, however, the seasonal variability seems not to explain the cycle. The model proposed in this article and the simulation-based analysis indicate that the spike separation is mainly controlled by the manner in which immunity wanes. Furthermore, simulations indicate that spikes predicted by the immunity dynamics may be significantly damped or be missing completely if the transmission rate drops, underlining the importance of mitigation measures. Simulations with a vaccination program included in the model also showed a similar effect. A missing spike, or several ones, may be accompanied by variability in spike separation times, thus suggesting that it may be difficult to retrieve information about the immunity dynamics by observing the spike separation alone.

According to the proposed model, the trend of the spike amplitudes depend on both the transmission rate and the vulnerability profile. Under the assumption of constant transmission rate, the spike train may repeat itself without attenuation, or the spike amplitudes may decrease. We observed that details of the vulnerability profile describing the immunity waning need not to be known to explain the trend, it is sufficient to have information about the robust statistics of the underlying fictitious probability measure that determines the profile.

In the post-pandemic era when the disease is becoming endemic and public health authorities have less tools to follow and control the spread, and reliable high-quality infection data becomes unattainable, fitting detailed parametrized models to data may be less relevant for disease prevention and control. Mathematical models continue to play an important role in identifying factors that could explain the observed infection patterns, thus helping identify robust features of available data that contain pertinent but non-detailed information of the state of the infection. Features that are can be presumed from sewage [18, 20, 21] and hospital data are the separation of the spikes and the trends in spike amplitude. The present contribution suggest how those features may be related to the infection dynamics.

## Data Availability

No data used or generated

## Acknowledgements

This work was partly supported by the NSF PIPP Phase I Award 2200255. In addition, the work of DC was partly supported by NSF DMS Award 1951446 and that of ES by NSF DMS Award 2204618.

